# Access to healthcare as an important moderating variable for understanding geography of immunity levels for COVID-19 - preliminary insights from Poland

**DOI:** 10.1101/2021.12.08.21267167

**Authors:** Andrzej Jarynowski, Vitaly Belik

## Abstract

**Background:** Biases in COVID-19 burden and uncertainty in estimation of the corresponding epidemiologic indexes is a known and common phenomenon in infectious diseases. We investigated to what extent healthcare access (HCA) related supply/demand interfered with registered data on COVID-19 in Poland.

**Material and methods:** We run a multiple linear regression model with interactions to explain geographic variation in seroprevalence, hospitalizations (on voivodeship – NUTS-2 level) and current (beginning of the 4th wave – 15.09-21.11.2021) case notifications/crude mortality (on poviat – old NUTS-4 level). We took vaccination coverage and cumulative case notifications up to the so called 3rd wave as predictor variables and supply/demand (HCA) as moderating variables.

**Results:** HCA with interacting terms (mainly demand) explained to the great extent the variance of current incidence and most variance of current mortality. HCA (mainly supply) is significantly moderating cumulative case notifications till the 3rd wave explaining the variance in seroprevalence and hospitalization.

**Conclusions:** Seeking causal relations between vaccination-or infection-gained immunity level and current infection dynamics could be misleading without understanding socio-epidemiologic context such as the moderating role of HCA (sensu lato). After quantification, HCA could be incorporated into epidemiologic models for improved prediction of real disease burden.

## Background

### HCA and infectious diseases

From epidemiologic practice, we already know that registered disease case notifications are not covering the real burden of infections to the same extent in various geographic locations due to socio-economic-medical inequalities [1]. Therefore, healthcare access (HCA) has already been identified as a possible factor influencing measurable COVID-19 epidemic indexes and explaining intra-country variation [2–5]. Thus, more resource-rich regions were able to organize both testing and treating capacities [6] and vaccination campaigns [7] in a faster and more inclusive way, whereas resource-poor regions were much more selective in the pandemic healthcare services delivery.

### Epidemiological modeling and healthcare analytics

Knowing immunity levels is important for proper prediction of the COVID-19 dynamics for a given region. Researchers proposed multiple methods and models combining natural immunity possibly acquired after infection with induced immunity by various vaccines taking into account their interactions as well as various immunity waning schema [8,9]. We do know quite well how various vaccines are waning efficacy [10] against symptomatic infections (controlled for covariates such as seasonality or virus variants), however post-infection immunity is much more difficult to assess [11]. Seroprevalence studies can be also used to estimate immunity levels, and have previously been used extensively to estimate infection fatality rate (IFR) [12,13]).

### Modeling Paradoxes

To unravel the so-called “epidemiological mystery” of COVID-19 [14] it is currently being attempted to identify the contextual factors associated with case and death reports with real infection and fatality rates. Researchers from Institute for Health Metrics and Evaluation [14] estimated that the cumulative number of unreported cases until the so-called 4th wave vary significantly even among countries within close geographic proximity. For instance for Eastern Europe, in Slovakia, reported cases should be increased by 13% corresponding to undiagnosed cases to reach the estimate, in Poland it’s 517%, while in Russia it’s 2230%. This example published in the Lancet reveals the scale of the problem associated with reporting and understanding COVID-19 outcomes. Moreover, recently some ecological studies are suggesting a weak or no link between vaccination coverage and current (Delta variant) epidemic dynamics in the general populations [15,16] as well as little to no effect of NPI (nonpharmaceutical Interventions) on COVID-19 incidence and mortality [17,18], which has gained a lot of controversy, and our observations put a new light on them.

### HCA and modeling

Surprisingly, HCA has not yet been significantly addressed in the practice of infection dynamics modeling [5]. While immunity is gained and lost at the individual level, immunity level often is tried to be quantified at a population level, for instance, by accounting for biased measures such as estimates of undiagnosed cases or eliciting seroprevalence via surveys. Wrong estimates of COVID-19 immunity levels varying over time and regions make predictions challenging and significantly reduce predictive power. Therefore, forecast results often deviate from reality, as was the case in Poland between June – September 2021 [19]. It is worth mentioning that a simple forecasting model “PL_GRedlarski-DistrictsSum’’ summing up extrapolation of COVID-19 cases’ trajectories in poviats (implicitly assuming their idiosyncrasy and heterogeneity driven by HCA among others) created by a volunteer from Medical University of Gdańsk is overperforming in short and medium term all other models [20], although the latter are financed and used by Polish Ministry of Health or ECDC. Albeit we could simply mechanistically mimic reporting patterns, we still do not know what are intra-country differences in shares of unreported cases. More or less accurate inter-country estimates in Eastern Europe (sharing the similar post-communistic social and health care patterns) are showing variability of over two orders of magnitude [14].

### Exploratory aim

We show on the example of Poland that **omitting HCA confounding/moderating factors could lead to misinterpretation in understanding current epidemic dynamics due to biased estimation of immunity levels and other epidemiologic indexes**.

## Material and methods

### Data description

Data obtained from Polish registries of 16 voivodeships (NUTS-2) and 380 poviats (old NUTS-4) only from publicly available various official sources or their archived versions. For instance, historical hospitalization rates are not publicly available and only can be scrapped from the archives of governmental webpages. We considered the following variables for our model.

### Independent variables

- **Cumulative No, cases per capita**: The cumulative numbers of COVID-19 notifications till the so-called 3rd wave of epidemic (04.03.2020 – 15.06.2021) for poviats and voivodeships [21] divided by its population size.
- **Fraction of vaccinated:** Percentage of vaccinated with at least one dose for poviats and voivodeships (at the end of 3^rd^ wave at 15.06.2021) for all age groups [22]. Vaccination coverage gives us a proxy of proportions of population which gain post-vaccination immunity before the so-called 4th wave.

### Moderating variables

- **Healthcare Access – Supply (supply HCA):** The number of physicians working in health care per 10,000 inhabitants in 2019 as an indicator of the supply HCA for poviat and voivodeship [23]. This is a good proxy for capacity and accessibility of healthcare (public and private).
- **Healthcare Access – Demand (demand HCA):** The number of consultations in primary care provided in 2019 for poviats and voivodeships as an indicator of demand HCA [23]. Pearson correlation between demand HCAs in 2019 and 2020 is 0.998 [7], so no significant regional changes have been observed in the demand for HCA due to the pandemic. Demand HCA is a complex conglomerate of attitudes towards healthcare (i.e., level of trust in the effectiveness of treatment offered by public healthcare), perceptions of accessibility (i.e., how easily one can reach healthcare facilities), and disease burden (i.e., elderly and inferior health populations are more likely to seek for healthcare).

### Dependent variables

- **Normalized incidence Sep/Oct’21**: 2-week incidence of COVID-19 notifications (21.09–04.10.2021) during the beginning of the so-called 4th epidemic wave for a poviat [21].
- **Normalized deaths Sep/Nov’21**: Crude mortality rate – Cumulative number of COVID-19 death cases (15.09–21.11.2021) during the so-called 4^th^ epidemic wave per poviat divided by its population size [24].
- **Normalized hospitalizations**: Number of occupied hospital beds (14.10.2021) at the beginning of the so-called 4th epidemic wave per voivodeship divided by its population size [25].
- **Obser-Co:** The fraction of seroconverted [26] per voivodeship in a random (by design) sample as a proxy for immunity level collected during 29.03–-14.05.2021. As this was the end of the 3rd epidemic wave, as well as the vaccination roll-out was at the beginning, this variable is a good proxy of post-infection acquired immunity.

The dynamics of the SARS-CoV-2 spread vary across the spatial clusters and initial conditions as number of index cases and immunity levels could lead to different phases at the beginning of each wave. We chose the beginning of the 4th wave for epidemiologic indexes (dependent variables): hospitalizations and incidence due to relatively equal distribution of Delta variants across country, when already the wealthier part of population returns from overseas vacations [27], children return to school but before the start of the new academic year (possible movement of population to large cities), in order to avoid some artefacts due to introduction or re-emergence of variants [6,28]. For mortality rates, we decided to take a longer period (up to maximum in incidence of 4th wave), in order to gather enough data.

### HCA access definition

Supply/demand HCA are assumed to be moderating variables that affects the relationship between independent variables (vaccine or post-infection immunity at the end of the 3^rd^ wave) and dependent variables (seroprevalence and the 4^th^ wave outbreak outcomes).

Our bidimensional HCA measure is a combination of [5,14,29–33]:

- Regional healthcare capacity or quality (corresponding to supply HCA) which other researchers have sought to examine, such as Health Security Index, Universal health coverage Index, Healthcare Accessibility and Quality Index, Human Development Index, number of physicians or nurses per capita, number of hospital beds per capita, government and total health spending per capita, number of tests performed per capita, Cumulative Standardized Testability Ratio;
- Behavioral patterns of using healthcare related to population demographic or comorbidity structure (corresponding to demand HCA) which other researchers were trying to capture by, for instance, age structure (e.g., share of seniors), BMI (e.g., share of obese), pulmonary or immunodeficiency diseases prevalence, number of medical consultations, share of smokers;
- The affective social conditions corresponding to the way how people are likely to seek healthcare services (corresponding to demand HCA) which other researchers have tried to capture by, for instance, magical thinking prevalence (e.g., interests in alternative medicine), ethnic structure, political populism, income inequality (e.g., intra-or inter-regional Gini coefficient), trust in medical professionals, government, or science, and affective autonomy (the normative tendency for people to maximize their own utility).

Thus, our socio-medical concept of HCA is covering a wide range of complex constructs and the goal of this paper is to demonstrate its importance.

### Regression models

We provide an empirical, rather than theoretical, approach for analyzing links between dependent and independent variables moderated by covariates (HCA). The common zero approach (but not state of the art [34]) for assessing a causal model is a multiple regression analysis [35] with interactions between variables (moderating and independent). Multiple linear regression was proposed in 4 equations n∈{1,2,3,4} with 4 predictors consisting of i∈{1,2} independent and i∈{3,4} moderating variables:

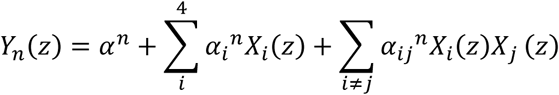

Where generally: Y_n_ are dependent variables; X_1_ and X_2_ are independent variables; X_3_ and X_4_ are moderating variables, X_i_X_j_ are 2-way interactions terms among predictors.

Where specifically: Y_1_ – seroprevalence – Obser-Co; Y_2_ – Normalized Hospitalizations; Y_3_ – Normalized incidence Sep/Oct’21; Y_4_ – Normalized deaths Sep/Nov’21; z is one of 18 voivodships for Y_1_ and Y_2_ or one of 380 poviats for Y_3_ and Y_4_; X_1_ – Cumulative No. cases per capita (independent variable); X_2_ – Fraction of vaccinated (independent variable); X_3_ – Healthcare Access – Supply/supply HCA (moderating variable); X_4_ – Healthcare Access – Demand/demand HCA (moderating variable); α are intercepts (without subscript indices) and slope parameters (with subscript indices).

Flow of explained variance of each dependent Y_n_ variable (in %) by predictors X_i_ and interactions among them is drawn on diagrams (Fig. 3-6). Models at the level of each geographical unit (poviat and voivodeship) should be compared separately. Because there is a known geographical phenomenon that the use of larger spatial units (NUTS-3, voivodeships) results in a better adjustment of statistical models (higher explained variance), due to the aggregation (averaging) of data for these units [36]. Graphical representation of possible causation links on diagrams is an informal visualization tool in our case, but has a formal mathematical form [37] for instance in Structural Equation Modeling (SEM), Path Diagrams or Directed Acyclic Graphs (DAG). Our model does not formally recognize independent or moderating variables, so presentation of directionality of interactions terms on diagrams is a priori convention only.

## Results

### Data exploration

Descriptive statistics of variables were analyzed. In a hierarchical arrangement (Fig. 1) of the correlation matrix and corresponding dendrograms, clusters of correlated related variables can be noticed.

**Fig. 1.**
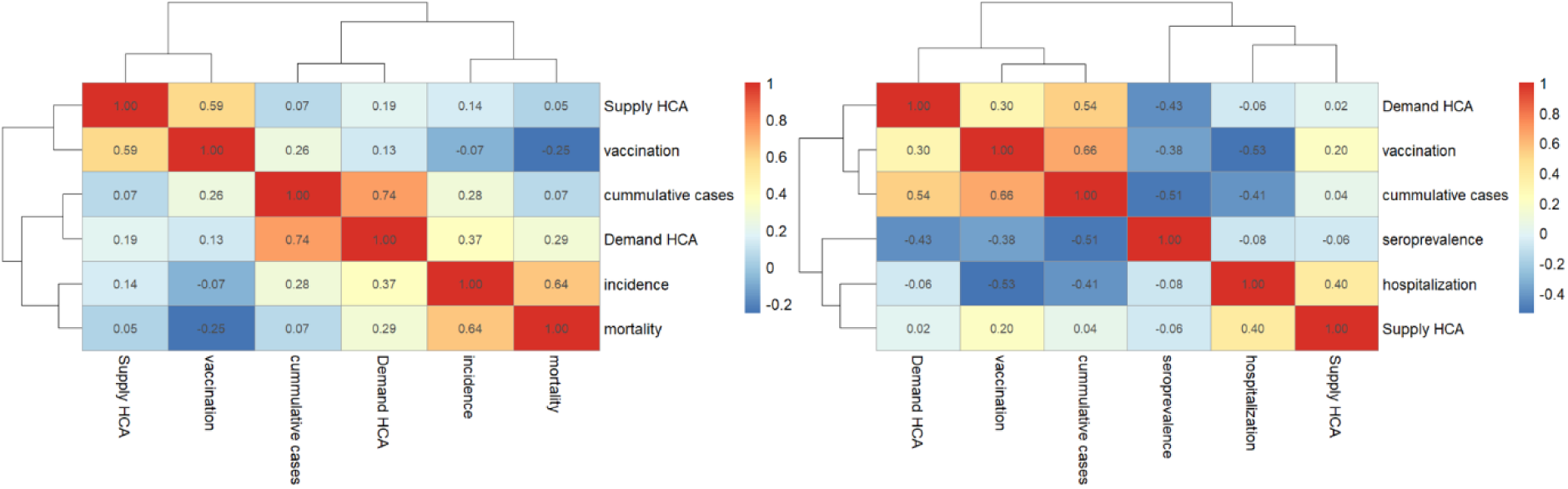

(Right) level. Independent and moderating variables available on poviat level have been aggregated to voivodeship level.

There is a very strong and significant (R=0.74, p-V=0.03) correlation between demand HCA and cumulative cases per capita (Fig. 1 Left). To remind, Demand HCA was assessed before the pandemic started, thus geographic distribution of reported normalized cases aggregated over the first three waves is highly dependent on the structural patterns of how people tend to use healthcare. On the other hand, supply HCA is strongly correlated with vaccination rate (R=0.59, p-V=0.09). This could mean that the vaccination program was more popular in regions with higher healthcare capacity. On voivodeship level (Fig. 1 Right) seroprevalence is significantly negatively correlated with Demand HCA (R=-0.43, p-V=0.05) as well as negatively correlated with cumulative cases (R=-0.51, p-V=0.06). It is important to note the paradox, that higher reported cumulative cases correspond to lower seroconversion rate, which can only be explained by structural geographical biases (for instance, due to intra-voivodeship variation in the variables being compared or inter-voivodeship variation of undiagnosed cases).

Unfortunately, hospitalization rates are not available on poviat level, so we choose to visualize geography of cumulative and current incidences mediated by Supply HCA (Fig. 2 left) and vaccination rates and crude mortality moderated by Demand HCA (Fig. 2 right).

**Fig. 2.**
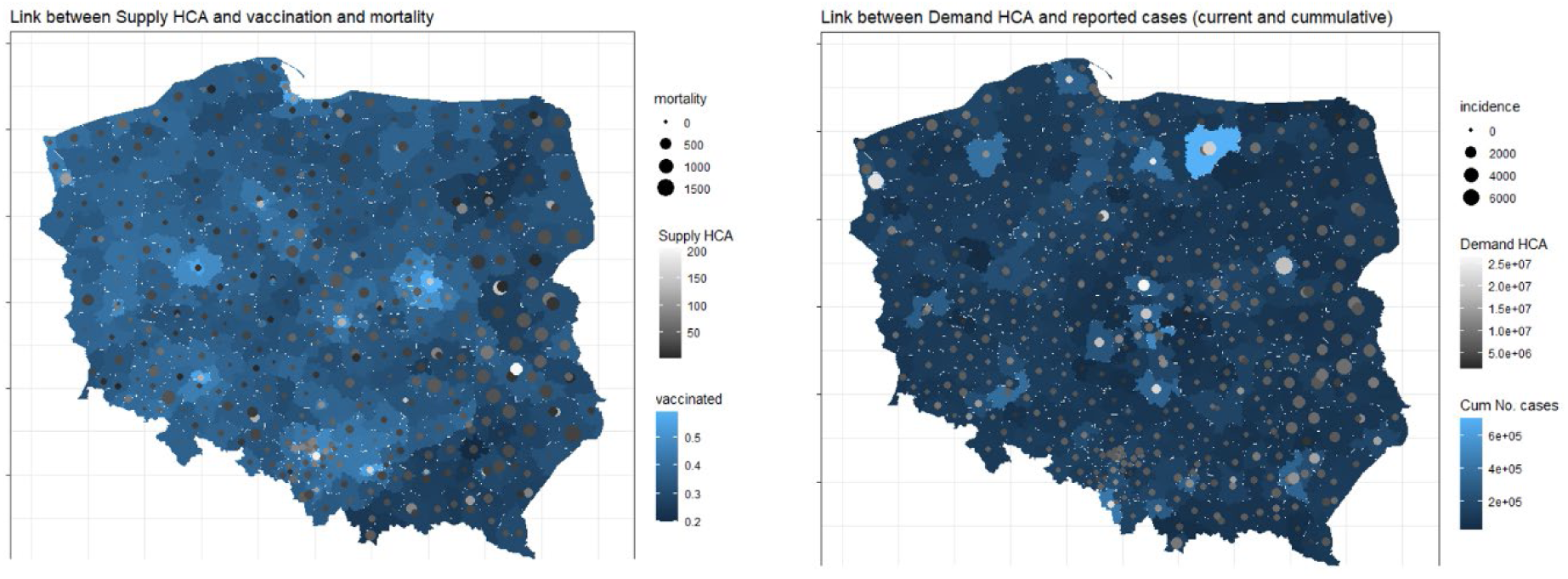
Map of the selected significant determinants of COVID-19 outcomes on poviat level. Left: with Supply HCA, Right: with Demand HCA.

Mapping (Fig. 2) shows that both moderating HCAs are highly granulated and do not cluster spatially to such extent as independent variables [6,7]. Thus, due to high heterogeneity within even close geographic neighborhoods (for instance between big cities and rural poviats), analysis on the level of voivodeship could be not sufficient.

### Regression models

The following predictor variables were used: vaccination coverage, cumulative case notifications, conditions moderating supply/demand HCA as well as 2-way interactions among them (Tab. 1, Fig. 3-6).

**Figure. 3.**
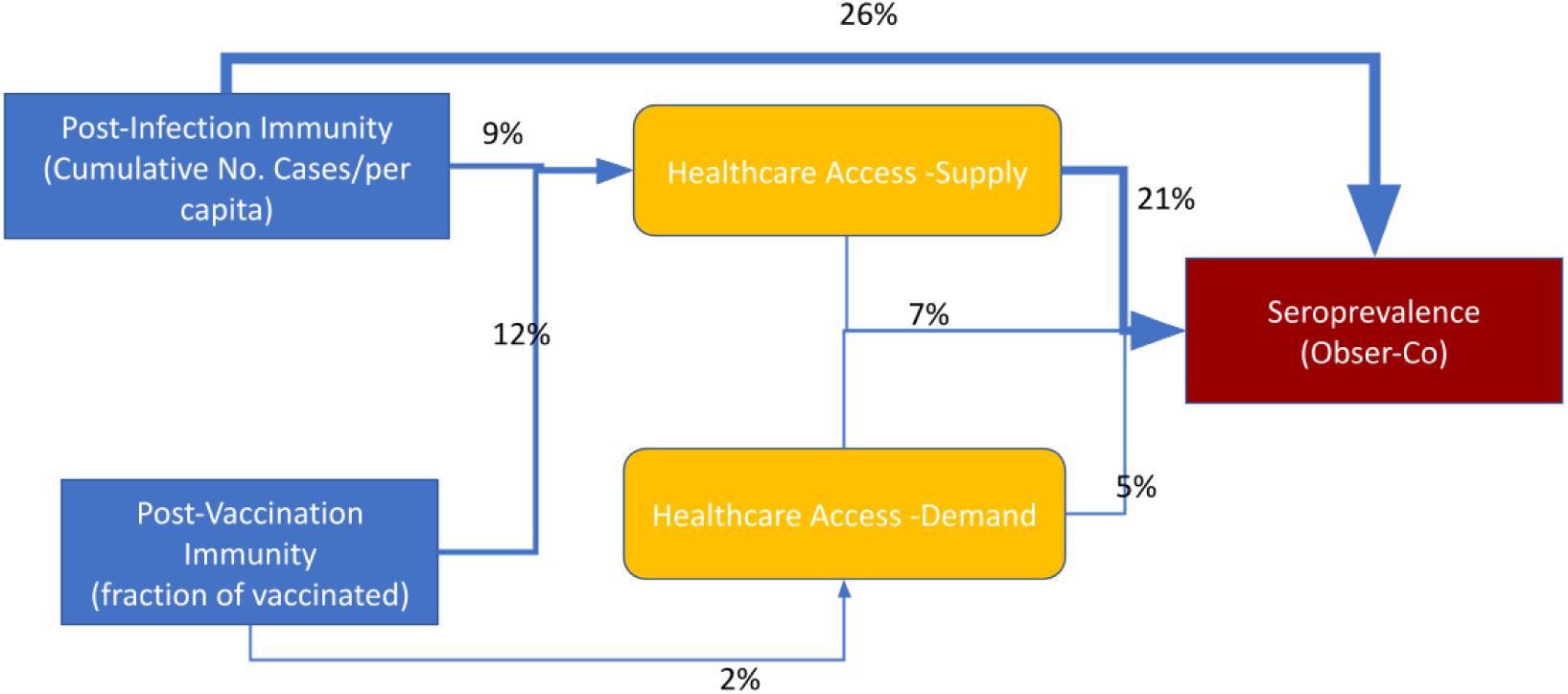
Explained variance of seroprevalence at the end of 3rd wave – immunity level estimation from Obser-Co [26] on voivodeship level.

The models presented in Table 1 were not selected according to statistical criteria. Therefore, there may be other models based on the same set of predicting variables with a better fit. However, these simple regressions allow us to highlight which variables and which interactions need further investigation. More variance is explained for hospitalizations (82%) than seroprevalence (61%) on voivodeship level. More variance is explained for deaths (25%) than incidence (23%) on poviat level.

**Table 1.** Percent of explained variance (%Exp) and significance level (p-V) for dependent variables (red) for given selection of predictors with interactions as independent variables (blue) or moderating variables (yellow).

| dependent variable | Seroprevalence |  | Hospitalizations |  | Incidence* |  | Deaths |  |
| --- | --- | --- | --- | --- | --- | --- | --- | --- |
| predictor | p-V | % Exp | p-V | % Exp | p-V | % Exp | p-V | %Exp |
| **cumulative cases** | 0.129 | 26 | 0.022 | 17 | 0.480 | 0 | 0.112 | 1 |
| **fraction of vaccinated** | 0.840 | 0 | 0.017 | 20 | 0.129 | 0 | <0.001 | 9 |
| **supply HCA** | 0.889 | 0 | 0.022 | 17 | <0.001 | 3 | 0.375 | 0 |
| **demand HCA** | 0.543 | 3 | 0.191 | 4 | <0.001 | 15 | <0.001 | 13 |
| **cumulative cases: supply HCA** | 0.330 | 9 | 0.719 | 0 | 0.006 | 1 | 0.172 | 0 |
| **cumulative cases: demand HCA** | 0.976 | 0 | 0.564 | 1 | 0.594 | 0 | 0.875 | 0 |
| **cumulative cases: fraction of vaccinated** | 0.900 | 0 | 0.027 | 15 | 0.003 | 3 | 0.012 | 1 |
| **demand HCA: fraction of vaccinated** | 0.611 | 2 | 0.549 | 1 | 0.016 | 1 | 0.007 | 1 |
| **supply HCA: fraction of vaccinated** | 0.266 | 12 | 0.038 | 12 | 0.461 | 0 | 0.364 | 0 |
| **supply HCA: demand HCA** | 0.393 | 7 | 0.118 | 6 | 0.068 | 1 | 0.336 | 0 |
| Residuals | - | 39 | - | 18 | - | 77 | - | 75 |

\* Incidence is sensitive to quadratic as well 3<sup>rd</sup> way interaction terms, thus it will need further robustness analysis

In a series of diagrams, we depict the explained share of variance for selected dependent variables of interest as calculated by multiple regression.

HCA (mainly supply indirectly) is significantly influencing seroprevalence survey results (Fig. 3). Thus, 9% of variance is explained by the interaction between cumulative case notifications and supply HCA. It’s worth mentioning that Demand HCA is even significantly correlated with seroprevalence (see. Fig. 1 left), which has not been replicated in the regression model (only 3% of explained variance directly). Most of the variance in seroprevalence is explained directly (26%) by cumulative cases notifications. Vaccination was in a very early stage at the time of seroprevalence survey, however there is an interaction pattern between supply HCA and vaccination (12%).

Hospitalizations seem to be well predicted by vaccination coverage and cumulative cases notifications (47% of variance considering also the interaction between them). HCA (mainly supply) affects hospitalizations (Fig. 4), but 12% of variance is explained by the interaction between vaccination coverage and supply HCA.

**Figure. 4.**
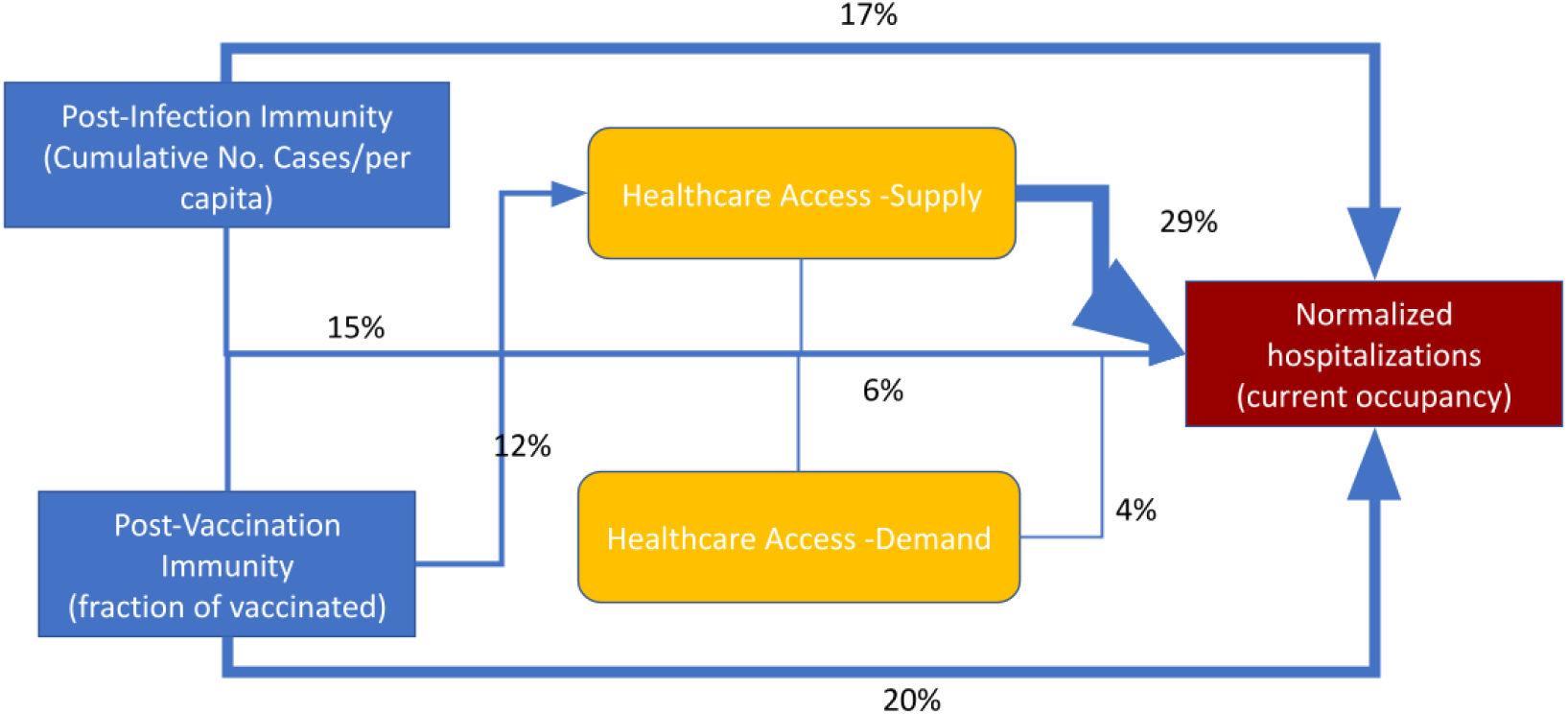
Explained variance of hospitalizations during the 4^th^ wave on voivodeship level.

HCA (mainly demand) is extremely important (Fig. 5, 6) to predict current infections dynamic (incidence and mortality). It is worth stressing that direct links between vaccination coverage as well as cumulative case notifications with current incidence are statistically negligible (only interactions with these terms have some impact).

**Figure. 5.**
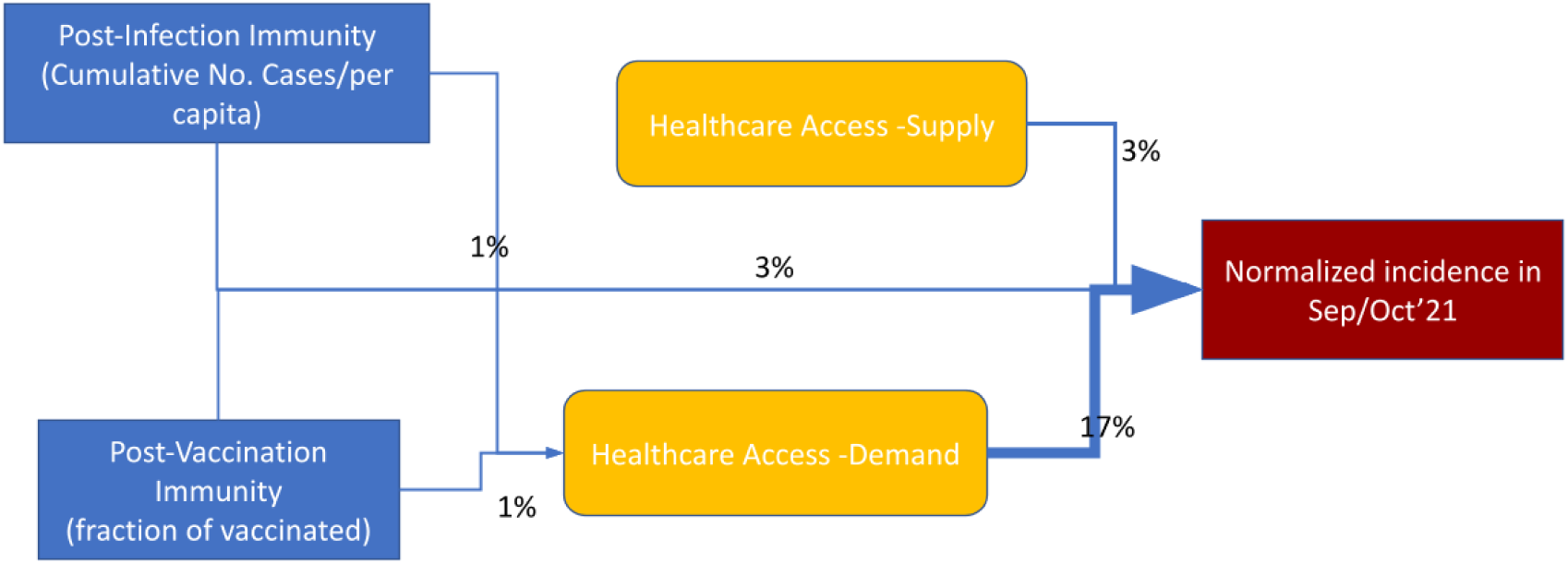
Explained variance of normalized case notifications (14-days incidence) during the 4^th^ wave on poviat level.

**Figure. 6.**
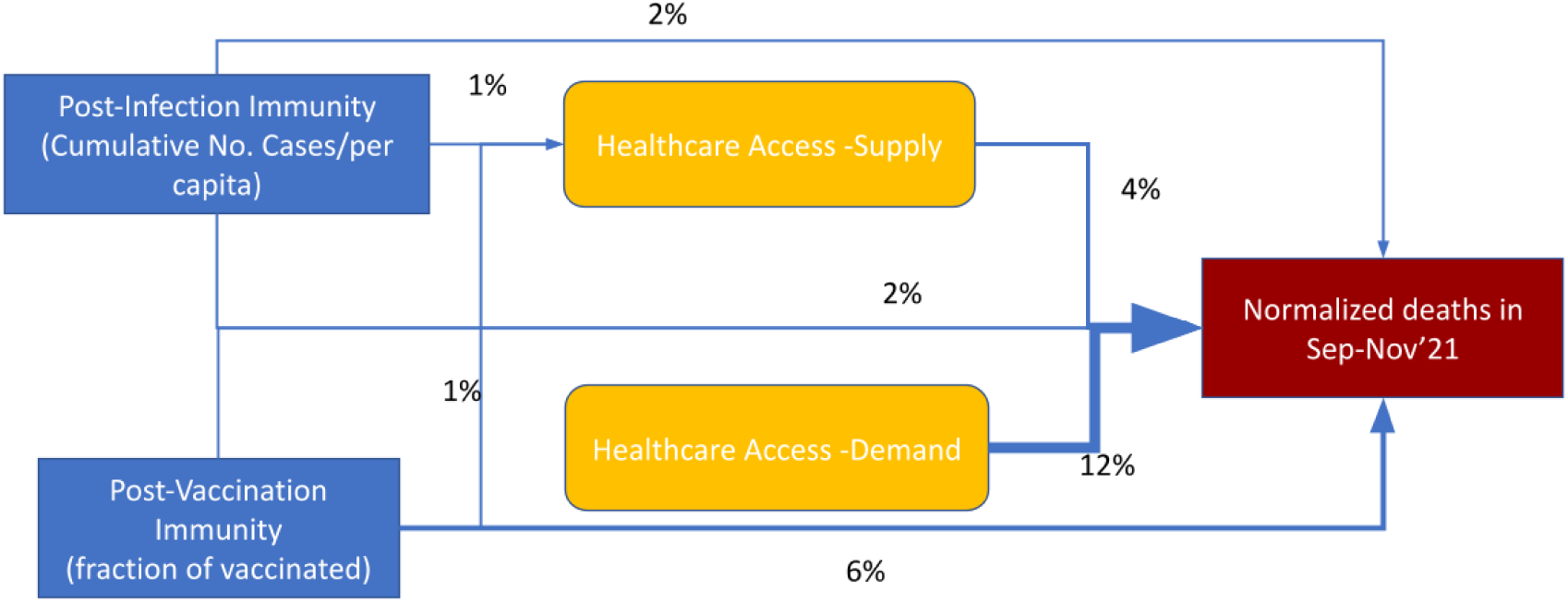
Explained variance of crude mortality rate during the 4^th^ wave on poviat level.

Crude mortality rate (Fig. 6) in comparison to incidence (Fig. 5) during the beginning of the 4th wave is less prone to the confounding effect of HCA. We can see a significant link (Fig. 6), between vaccination coverage and crude mortality rate due to COVID-19.

## Conclusions

### Discussion of regressions

There are multiple possible mechanisms explaining mediating role of HCA (however causal statements cannot be defined as probably both explaining and explanatory variable seems to depend on HCA):

- In regions with high healthcare service/worker availability (supply HCA) patients are more likely to be sampled as seroconverted (Fig. 3), have a higher chance to find a hospital bed (Fig. 4), as well as the chance of being tested (and get positive result) is increased (Fig. 5).
- Case notifications, as well as in some extent crude mortality (Fig. 5, 6), are highly correlated with the way how patients are likely to use the public healthcare system (demand HCA), thus in regions with low demand HCA real number of infections seems to be underestimated by documented case notifications to a higher extent than in places with high demand HCA.

We found that hospitalizations seem to be the most understandable for a given set of predictor variables (the highest variance explained and potentially strong model robustness) as well the least dependent on HCA (Tab. 1, Fig. 4). On the other hand, incidence is the most difficult to assess and interpret (the lowest variance explained and weak model robustness) as well as the most prone to be driven by HCA (Tab. 1, Fig. 5).

### Modeling paradoxes – interpretation

Concluding, each epidemiological index is more or less confounded by HCA in different ways: seroprevalence obtained via surveys and hospitalizations mainly by supply HCA, but documented case notifications and crude mortality rates by mainly demand HCA. As all of investigated epidemiologic indexes (Tab. 1) are highly dependent on HCA within a single country with similar surveillance methodology across regions (at least in theory), so inter-country comparison or measuring effectiveness of vaccination or NPIs for a selected index lead to possible biases in interpretation [14,16–18] even unintentionally. As we found that reported geographic variability of incidence is more prone to be driven by socio-epidemiological factors rather than infectious dynamic process, all ecological analysis using this outcome must be interpreted with extreme caution. Moreover, the lack of or even slightly positive correlations between reported incidence (during the 3rd and 4th wave) and vaccination rates by regions has been confirmed in our study to be significantly confounded by HCA (Fig. 5). As this argument is often used by the so-called anti-vaccination/anti-sanitarian movements, it is worth mentioning that causal relation cannot be claimed with the observed moderating role of HCA (possible due to bias in tendency of seeking available healthcare in the mild course of the disease). Thus, vaccination was not only averting hospitalizations [25] or death tolls [38] in Poland (Fig. 4, 6), but also probably has reduced transmission probability (at least until introduction of Omicron variant), although it may be partially masked by inequalities in HCA (Tab. 1, Fig. 5).

### Limitation and future works

It’s important to mention that regression with incidence as an independent variable is the least robust (sensitive to adding 3-way interaction terms as well nonlinear forms of dependent variables), however this is out of scope of this preliminary work. This study is only an exploratory, so-called “zero” approach to illustrate the mediating phenomenon due HCA differences between geographical regions. Our approach has multiple limitations because vaccination coverage, cumulative case notifications, seroprevalence, and current outbreak dynamics are snapshotted at a given time point and were considered for different time periods. Due to availability of explained variables on different levels (voivodeship or poviat) model comparison should be done with caution. Other possible interfering variables were not taken into consideration (such as socio-economic-demographic portrait of the population) and other studies found that HCA could be collinear with multiple variables [39]. Further causal modeling using a longitudinal approach is required to support our preliminary observations. These results are specific to the Polish population / healthcare system and the role of the access to healthcare could be different in other settings.

### Recommendation

Moderating roles of HCA could be changing in time. However, this simple analysis suggests that including HCA indexes into models of disease dynamics will increase their short-term (meteorological conditions could be added as well [40]) and could increase long-term predictive power. Only triangulated pictures based on reported cases (with estimates of undiagnosed infections), deaths (due to COVID-19 and excess mortality), and hospitalizations (separately with and due to COVID-19), may allow for epidemiological interpretation. Thus, there are mathematical approaches of reducing the bias by combining multiple epidemiological indexes or constituting their derivatives [41,42]. For instance, crude fatality rates [43] seem to be less dependent on HCA than estimates of IFR in Poland. Moreover, recent suggestions of ECDC [44] and WHO [45] to put more emphasis on hospitalizations/mortality rather than laboratory confirmed cases in understanding the burden of disease are also supported by our findings. We regret that data gathered by state is not provided in an easy to analyze format [46] and a massive manual work is needed for data preparation. The most important and probably the least biased variable (according to our preliminary analysis) – hospitalization (being a good proxy for the severe cases) – is not available on poviat level (old NUTS-4) in Poland. We suggest that the number of hospitalized patients originating from a given region (poviat or even municipality) could be a very significant epidemiologic index.

## Data Availability

All data produced and processing scripts are available online at Github

https://github.com/ajarynowski/Healtcare_Access_COVID/

## Acknowledgment

Data, scripts in R and additional diagnostics are available on https://github.com/ajarynowski/Healtcare_Access_COVID. We thank Monika Wójta-Kempa, Magdalena Rosińska, Kamil Rakocy and Members of Polish Vaccinological Society for consultations. Study was partially supported by DFG (German Research Foundation, project number 458528774).

## Notes

### Competing Interest Statement

The authors have declared no competing interest.

### Funding Statement

This study was partially supported by DFG (German Research Foundation, project number 458528774).

### Author Declarations

Data:seroprevalence, hospitalization, case notifications, crude mortality, vaccination coverage, supply/demand healthcare access indexes. Sources: Statistics Poland, Ministry of Health Poland, Crisis Management Poland

### Summary of Updates

added fig. 1 and fig. 2 reorganized structure, reference updated

